# Spatial heterogeneity of T cell repertoire across NSCLC tumors, tumor edges, adjacent and distant lung tissues

**DOI:** 10.1101/2022.10.14.22281040

**Authors:** Qikang Hu, Yang Gao, Meredith Frank, Liyan Ji, Muyun Peng, Chen Chen, Bin Wang, Yan Hu, Zeyu Wu, Jina Li, Lu Shu, Qiongzhi He, Yingqian Zhang, Xuefeng Xia, Jianjun Zhang, Xin Yi, Alexandre Reuben, Fenglei Yu

**Affiliations:** Department of Thoracic Surgery, The Second Xiangya Hospital of Central South University, Changsha, P. R. China; Hunan Key Laboratory of Early Diagnosis and Precise Treatment of Lung Cancer, The Second Xiangya Hospital of Central South University, Changsha, China; Department of Thoracic Surgery, Xiangya Hospital, Central South University, Changsha, P. R. China; Early-Stage Lung Cancer Center, The Second Xiangya Hospital of Central South University, Changsha, China; Xiangya Lung Cancer Center, Xiangya Hospital, Central South University, Changsha, China; Hunan Engineering Research Center for Pulmonary Nodules Precise Diagnosis & Treatment, Changsha, China; National Clinical Research Center for Geriatric Disorders, Changsha, China; Department of Thoracic/Head and Neck Medical Oncology, University of Texas MD Anderson Cancer Center, Houston/United States of America; Geneplus-Beijing Institute, Beijing/China

**Keywords:** T cell repertoire, NSCLC, IHC

## Abstract

**Background:** A better understanding of the T cells in lung cancer and their distribution across tumor-adjacent lungs and the peripheral blood is needed to improve efficacy and minimize toxicity from immunotherapy to lung cancer patients.

**Methods:** Here, we performed CDR3β TCR sequencing of 143 samples from 21 patients with early-stage NSCLC including peripheral blood mononuclear cells, tumor, tumor edges (<1cm from tumor), as well as adjacent lungs 1cm, 2cm, 5cm, and 10cm away from the tumor to gain insight into the spatial heterogeneity of T cells across the lungs in patients with NSCLC. PD-L1, CD4 and CD8 expression was assessed by immunohistochemical staining and genomic features were derived by targeted sequencing of 1,021 cancer related genes.

**Results:** Our study reveals a decreasing gradient in TIL homology with the tumor-edge, adjacent lungs, and peripheral blood but no discernible distance-associated patterns of T cell trafficking within the adjacent lung itself. Furthermore, we show a decrease in pathogen-specific TCRs in regions with high T cell clonality and PD-L1 expression.

**Conclusions:** The exclusion in T cells at play across the lungs of patients with NSCLC may be potentially the mechanism for lung cancer occurrence.

## Introduction

Lung cancer is the leading cause of cancer-related deaths and is expected to claim over 130,000 lives in the US in 2022 alone[1] (https://www.cancer.org/research/cancer-facts-statistics/all-cancer-facts-figures/cancer-facts-figures-2022.html). About 85% of lung cancer diagnoses are classified as non-small cell lung cancer (NSCLC)[1, 2]. While treatments for early-stage NSCLC bode decent success rates, 75% of patients present with late-stage diseases at the time of diagnosis, for which survival rates are poor[2, 3]. Immunotherapies, such as immune checkpoint blockade (ICB) or adoptive cell therapy (ACT) using autologous T cells, have led to substantial clinical benefit, yet a majority of patients do not respond to treatment or develop secondary resistance[4, 5]. On the other hand, although it is overall better tolerated than conventional chemotherapy, ICB can lead to serious toxicities, some of which can be lethal. These have prompted exploration into immune-related drivers of suboptimal responses and toxicities to identify biomarkers to stratify patients for more personalized treatments.

PD-L1 is a widely used predictive marker. However, application across large patient datasets has resulted in inconsistent predictive power[6]. Tumor mutational burden (TMB) is another biomarker associated with efficacious responses to ICB and adoptive transfer of expanded autologous CD8^+^ T cells[7, 8]. NSCLC tumors with higher TMB generally have better clinical responses to ICB, perhaps due to a larger pool of neoantigen targets for CD8^+^ T cell recognition[6, 9-11].

CD8^+^ T cells are critical mediators of anti-tumor responses as multiple groups have shown that higher numbers of CD8^+^ tumor infiltrating lymphocytes (TIL) are associated with improved outcomes[12, 13]. More specifically, recent work from our group identified intratumor differences in the T cell repertoire as prognostic tools in NSCLC. Multi-region TCR sequencing revealed that greater TCR ITH was associated with greater risk of relapse and higher intratumor ITH in clonality was associated with more aggressive disease progression and greater risk of relapse[4]. Our more recent work demonstrated a high proportion of TCR overlap between the tumor and adjacent healthy lung tissue and greater TCR overlap was associated with worse survival[14]. In these studies, we determined that by analyzing the homology in T cell repertoire between tumors and their adjacent lungs, we could identify T cells more likely to recognize viral antigens (bystander T cells), and that these bystander T cells were associated with worse outcome. However, in our prior studies, we were not able to assess any spatial differences in the T cell repertoire of tumor-adjacent lungs based on proximity to the tumor[14]. Here, we used a step-wise approach to deconstruct the T cell repertoire architecture across 6 regions within NSCLC tumor tissue and the surrounding healthy lung tissue. T cell markers and repertoire metrics were compared across resected tissue from the tumor, tumor edge (<1cm from tumor), and 1, 2, 5, and 10 cm away from the tumor across 21 lung adenocarcinoma patients in order to better understand the impact of the tumor on the T cell repertoire across the lungs.

## Materials and Methods

### Tissue collection

Tissue samples from a total of 21 patients with primary lung cancer were collected at Second Xiangya Hospital of Central South University from September to December 2018. All patients gave written informed consent. The study was approved by the Ethics Committee of Second Xiangya Hospital Central South University (IRB: 2020084). Tissues from tumor, tumor edge (defined as <1 cm away from tumor), 1 cm away from tumor, 2 cm away from tumor, 5 cm away from tumor, 10 cm away from tumor were collected for each patient during resection.

### Targeted sequencing

DNA was extracted from FFPE of tumor tissues using Promega Maxwell™ RSC DNA FFPE Kit (Lot: AS1135#847221). Blood DNA was used as control. DNA (0.8-1.0 μg) was sheared into fragments with a peak of 200-250 bp for library preparation using NEBNext® Ultra™ DNA Library Prep Kit (NEB, Ipswich, MA). The barcoded libraries were captured by a customized panel of 1021 genes as previously described[15]. Sequencing was performed on a GeneSeq2000 (Suzhou GenePlus Clinical Laboratory Co, Suzhou, China) platform. Reads with low-quality (a. read with a half bases with quality ≤ 5; b. reads with N base ≥ 5%; c. reads with average base quality < 0) were removed from raw sequencing data. Then clean reads were mapped into hg19 human genome using bwa, and reads were further analyzed through sentieon pipeline. Somatic single nucleotide variations and small indels were called by MuTect2 and TNscope, respectively. Final SNVs and indels were filtered by variant allele frequency ≥ 1%.

### T cell receptor sequencing

Sequencing for human TCRβ chain complementarities determining region 3 (CDR3) was performed as previously described[16]. Briefly, V and J genes of CDR3 gDNA were amplified with multi-plex primers. The PCR products were sequenced after fragment selection by Illumina platform with paired-end 100bp. The clean data were obtained by removal of low-quality reads. Paired-end reads were used for MIXCR to map into V and J genes and annotated using ImMunoGeneTics (IMGT) database. Pathogen associated TCRs were clustered by GLIPH2[17] using pathogen-related Mc-PAS dataset.

### Immunohistochemistry

Tumor tissue PD-L1 was stained by PD-L1 (SP263) antibody (Roche). Experiments were performed as manufacture’s instruction. Positive PD-L1 staining defined as 1) >25% of tumor cells exhibit positive membrane staining 2) immune cells present (ICP) >1% and IC positive >=25% or 3) ICP = 1% and IC^+^ = 100%. The IHC results for PD-L1 were viewed by independent two pathologists and averaged together for analysis. CD4^-^ and CD8^-^ antibodies were provided from Servicebio Inc (Wuhan, China). Immunohistochemistry score of CD4 and CD8 positive staining were quantified by artificial intelligence-assistant IHC scoring system (Servicebio Inc, Wuhan, China).

### Statistical analysis

All the statistics and graphs were analyzed by R (v 4.1.0). The Mann-Whitney U test was used to determine the differences of numerous continuous data between groups. Correlation was performed by Spearman coefficiency. Paired t-tests and 2-way ANOVA were used when appropriate. Significant differences were considered if *p* < 0.05.

## Results

### Study design and patient cohort

To investigate the spatial T cell composition of the tumor and adjacent lung microenvironments, we enrolled a cohort of 21 patients with primary lung tumors (**Fig. 1A-B**). Patients had early-stage (stage I-III) NSCLC and underwent lobectomies for curative intent. Targeted sequencing was performed to evaluate the genomic landscape, and revealed a high prevalence of *TP53* mutations (55%, 11/20 patients), *EGFR* mutations (45%, 9/20 patients), and ZFHX3 (20%, 4/20 patients) among others (**Fig. 1C**). No differences in T cell repertoire were observed based on mutational landscape nor smoking status (**Supplementary Fig. 1-2**). Clonality and diversity were compared across all tissues for smokers versus non-smokers with no differences seen between the two groups across all tissue samples (**Supplementary Fig. 2**).

**Figure 1.**
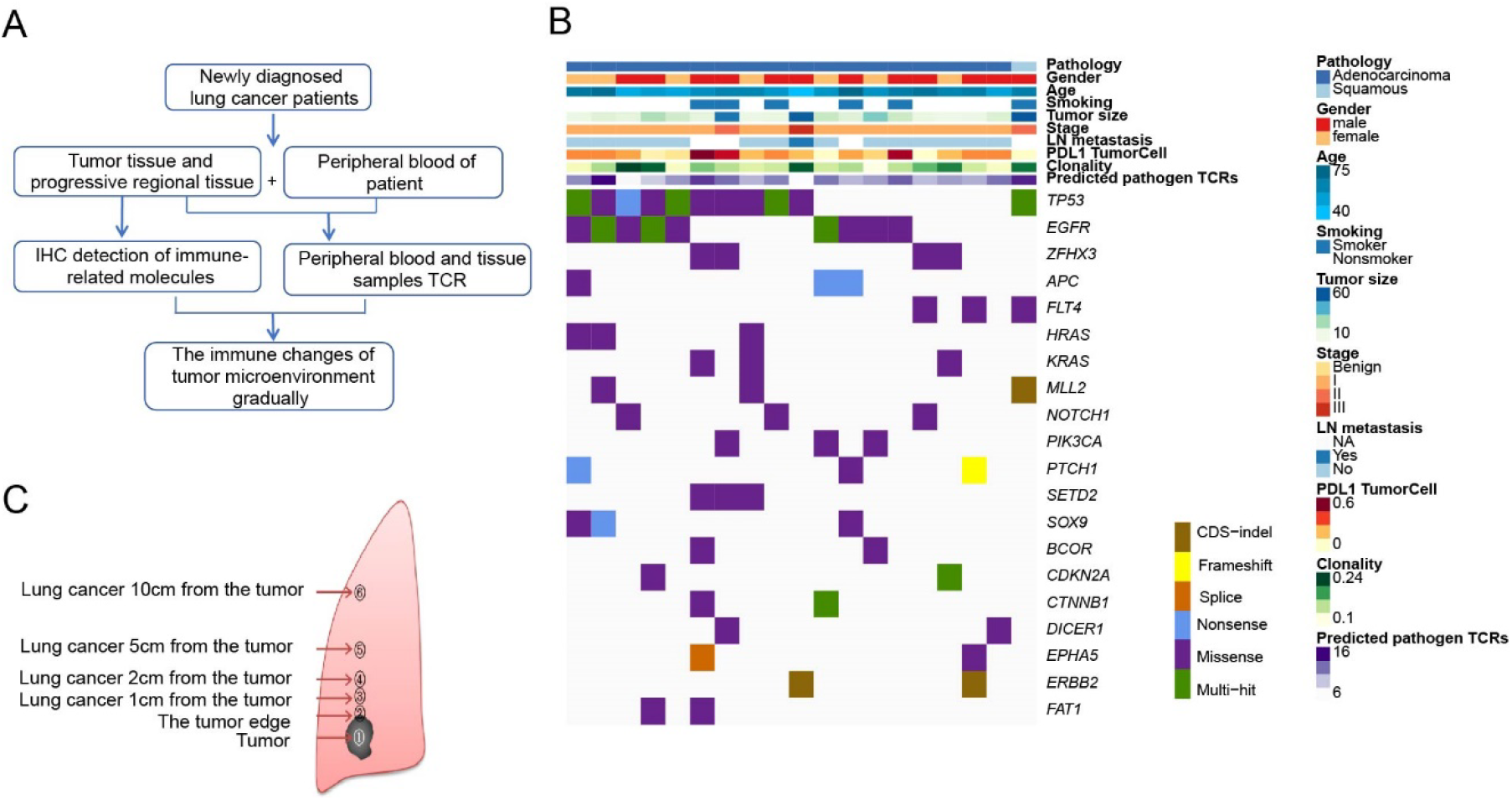
A: Schematic representation of experiment design. B: Tissue samples from a total of 6 regions were analyzed for TCR repertoire metrics across n=21 patients. C: Heatmap of clinical characteristics and tumor mutational data of top 20 mutated genes across n=21 patients.

### Increasing gradient of CD8+ and PD-L1+ observed in tumor periphery

Previous studies in NSCLC have demonstrated that greater CD8^+^ T cell densities within the tumor and tumor edge are associated with increased overall survival and, conversely, higher CD4+ T cell densities are associated with worsened survival [12, 13]. Accordingly, we assessed changes in CD8+ and CD4+ T cell densities within tumor tissue and peripheral regions by immunohistochemistry. No significant difference in CD8^+^ or CD4^+^ T cell densities was observed between tumor tissue and peripheral regions (**Fig. 2A-D**). However, the CD8^+^:CD4^+^ ratio was higher in the tumor edge, when compared with the tumor itself, indicating the presence of CD8^+^-rich regions in the tumor periphery (**Fig. 2E**). PD-L1 expression was also measured by immunohistochemistry but revealed no differences between regions. However, overall tumor peripheral regions exhibited higher PD-L1 expression on immune cells when compared to the tumor itself (**Fig. 2F-G**).

**Figure 2.**
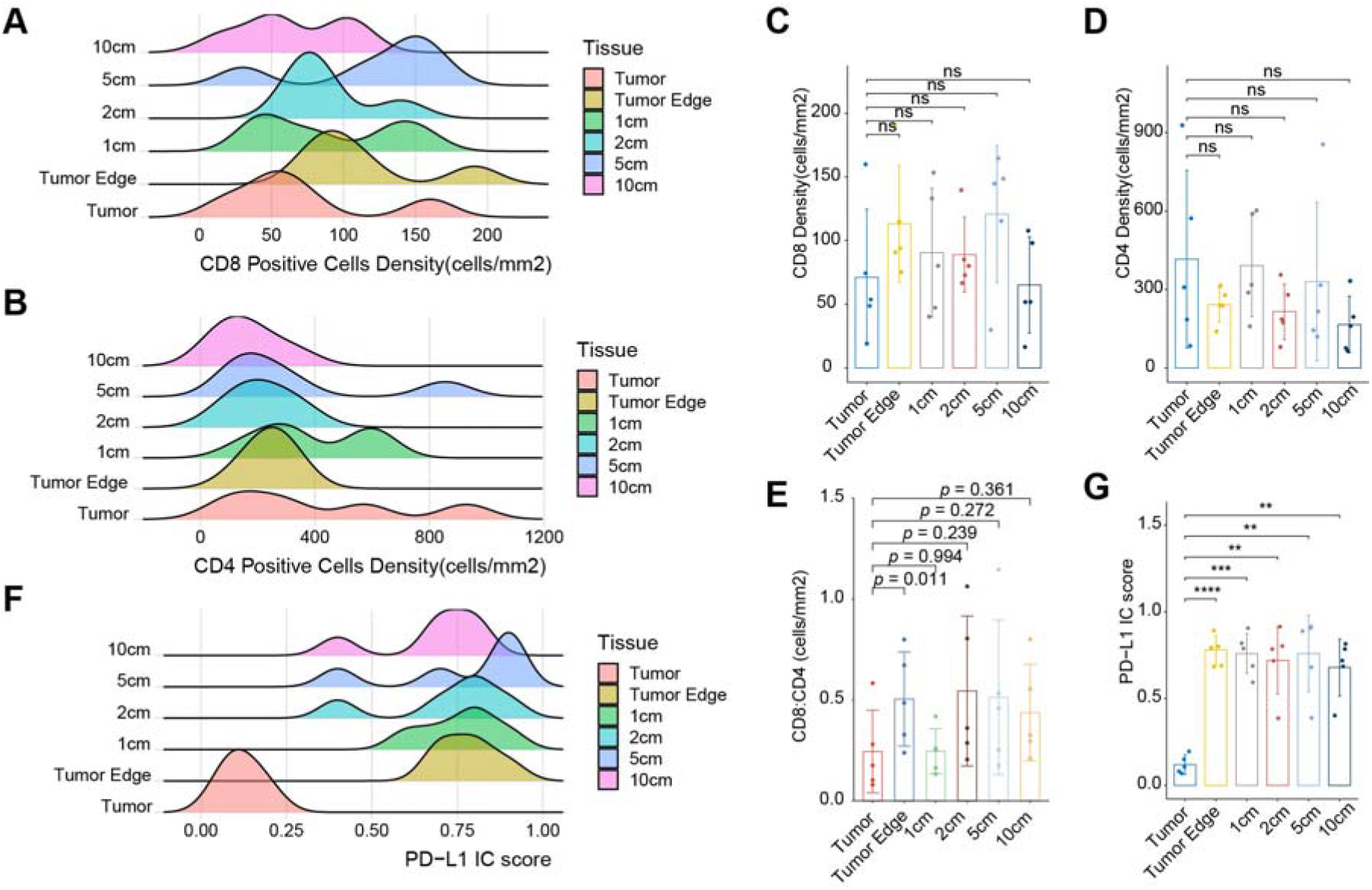
A: CD8+ cell density (cells/mm2) measured by IHC across all tissues for n=5 patients. B: CD4+ cell density (cells/mm2) measured by IHC across all tissues for n=5 patients. C: PD-L1 score measured by IHC across all tissues for n=5 patients. D: Median CD8+: CD4+ cell densities (cells/mm2) across all regions for n=5 patients.

### Regions of T cell high diversity exhibited lowest clonality and PD-L1 expression

To assess the spatial distribution of T cell repertoire, we performed sequencing of the CDR3β region of the T cell receptor primarily involved in antigen binding and analyzed related TCR metrics, namely, clonality and diversity. Clonality was calculated across regions for 21 patients (**Fig. 3A**). Tumor tissue exhibited the lowest level of clonality but highest level of diversity, whereas peripheral regions, namely 2 cm *(p= 4*.*9*^*e-7*^), contained higher clonality but lower amounts of diversity, consistent with prior studies by our group[14] and suggestive of a suppressed T cell repertoire within the tumor microenvironment *(p<0*.*05)* (**Fig. 3B-C**). PD-L1 expression was moderately associated with clonality *(rho= 0*.*43, p= 0*.*0178)*, namely at 1 cm away from the tumor (*R= 0*.*62, p= 0*.*26)* (**Fig. 3D-E**). No significant correlation was observed between clonality and CD8^+^ density across all tissues (**Supplementary Fig. 3**).

**Figure 3.**
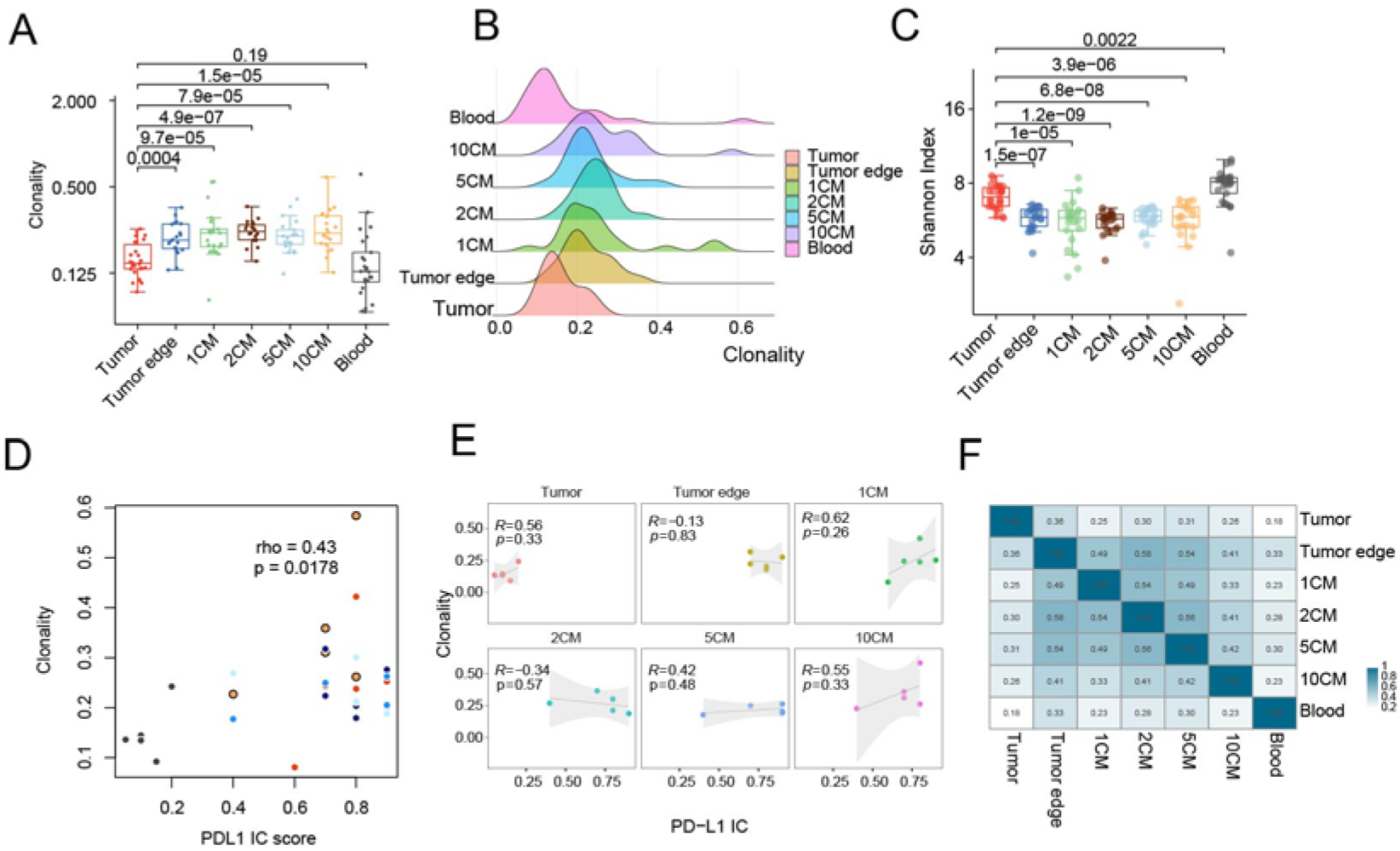
A: Morisita overlap values were calculated between tumor and normal tissue for n=21 patients. From top to bottom: tumor, tumor edge, 1cm, 2cm, 5cm, 10cm from tumor, peripheral blood, respectively. From left to right: tumor, tumor edge, 1cm, 2cm, 5cm, 10cm from tumor, peripheral blood. In most patients, the greatest amount of overlap was seen between normal tissues peripheral to the tumor, namely, the tumor edge, 1cm, 2cm, and 5cm. The lowest amount of overlap was seen between tumor tissue and remaining tissues. 3B: Average morisita overlap between regions for n=21 patients. From top to bottom: tumor, tumor edge, 1cm, 2cm, 5cm, 10cm from tumor, peripheral blood, respectively. From left to right: tumor, tumor edge, 1cm, 2cm, 5cm, 10cm from tumor, peripheral blood. 3C: Clonality values for all samples for n=21 patients. 3D: Median clonality across regions for n=17 patients. 3E: Median Shannon index values across regions for n=17 patients. 3F: Clonality vs PD-L1 expression (all tissues combined). Pearson’s coefficient was used to analyze the association between clonality and PD-L1 expression across all tissues for n=5 patients.3G: Clonality vs PD-L1 expression (stratified by tissues). Pearson’s coefficient was used to analyze the association between clonality and PD-L1 expression across all tissues for n=5 patients.

### Dominant T cell populations are better conserved in tumor margins compared to inside the tumors

Morisita overlap index (MOI) values were calculated between all available regions for 21 patients. As shown in **Supplementary Fig. 4**, substantial interpatient heterogeneity was observed among samples. As expected, lower amount of overlap was seen between tumor tissue and all other tissues, with a modest overlap only with the tumor edge (**Fig. 3F**) (MOI=0.36). Most patients exhibited the highest overlap between 2cm and surrounding regions, namely 5cm, 1cm and the tumor edge (**Fig. 3F**) (MOI=0.56, 0.54, and 0.58 respectively). In general, dominant T cell populations were better conserved between the tumor margins and “hot regions” of increased clonality compared to the tumors (**Fig. 3F**).

### High clonality regions are infiltrated by fewer predicted pathogen-specific T cells

As large numbers of bystander T cells in tumor tissue have been identified in NSCLC and other solid tumors[14, 18, 19], we next analyzed the pathogen-specificity within tissue regions. GLIPH analysis was used to cluster similar CDR3 motifs which were cross-referenced against publicly available viral CDR3 motifs to predict viral antigen-specificity. Predicted pathogen TCR counts were calculated for all regions across 21 patients (**Fig. 4A**). Pathogen-specific TCR counts varied significantly between regions with tumor tissue containing the highest proportion of pathogen-specific TCRs (**Fig. 4B**). Clonality was inversely proportional to pathogen count *(R=-0*.*3, p= 0*.*00081*, **Fig. 4C and Supplementary Fig. 5**). Interestingly, 2cm away from the tumor contained the highest clonality and also exhibited a strong negative association between amount of predicted pathogen TCRs and CD8^+^ density *(R=-0*.*87, p= 0*.*054)*, but not CD4^+^ cell density *(R=0*.*58, p= 0*.*3)*, potentially suggesting preferential expansion of non-viral CD8^+^ T cells close to tumor edge (**Supplementary Fig. 6**). In tumor tissue, no correlation was observed between CD4^+^ T cell density and the amount of predicted pathogen TCRs (**Fig. 4D**). In tumor tissues, the amount of pathogen-specific TCRs negatively correlated with TCR clonality (**Fig. 4E**) but positively with CD8^+^ T cell density (**Fig. 4F**) indicating the presence, but lack of expansion, of pathogen-specific T cells within the tumor (**Fig. 4F**). A moderate, but not statistically-significant, association was observed between the proportion of predicted pathogen TCRs and tumor cell PD-L1 expression *(R= 0*.*43, p=0*.*059)* (**Fig. 4G**).

**Figure 4.**
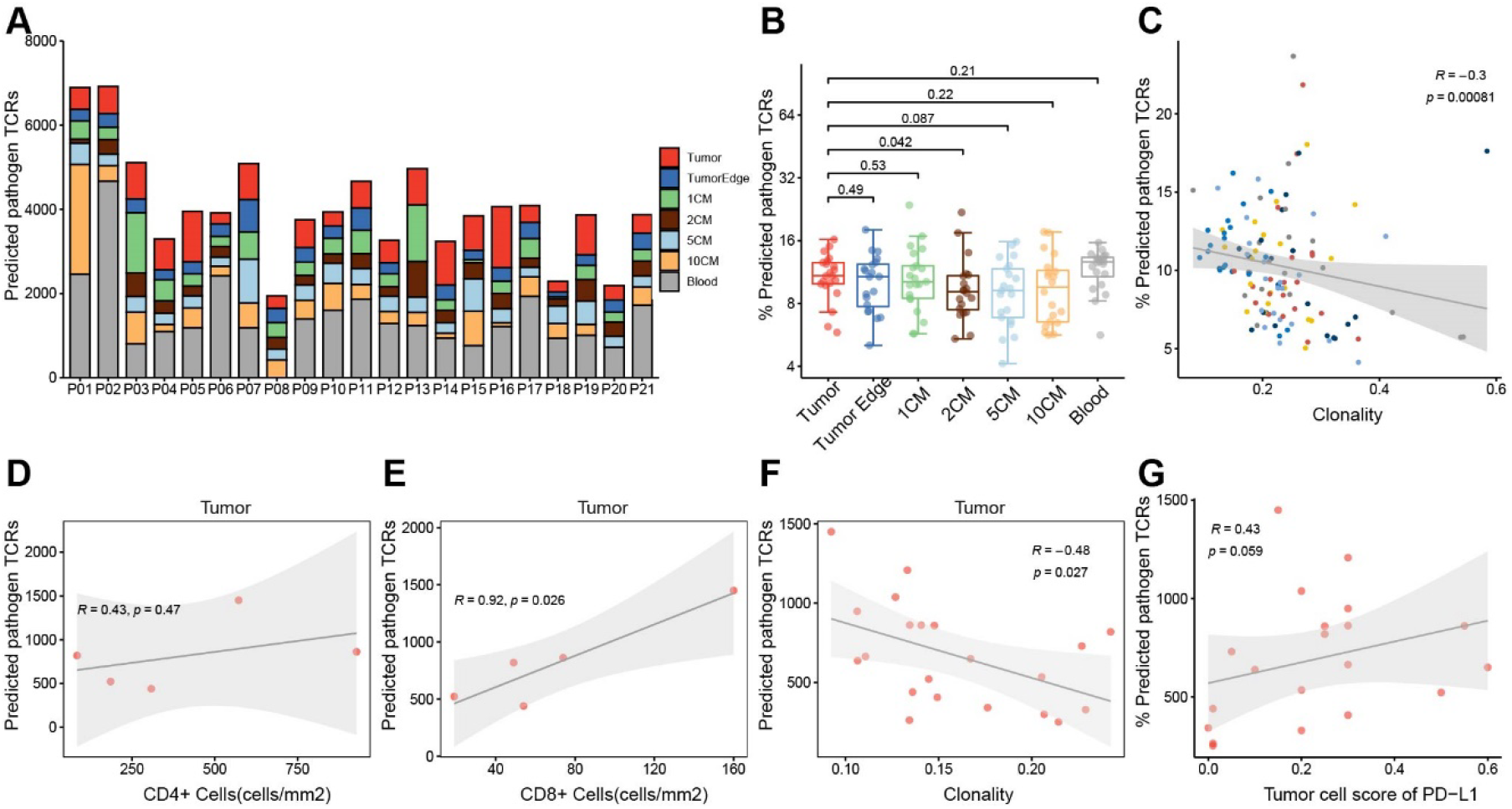
A: Percentage of predicted pathogen TCRs across all regions for n=21 patients. 4B: Median percentage of predicted pathogen TCRs across all regions for n=21 patients.4C: Regional clonality values were plotted against median proportion of predicted pathogen TCRs for all tissues. The solid line represents correlation between clonality and predicted pathogen TCRs. Thin dotted lines represent the 95% confidence interval. 4D: Predicted pathogen TCRs (absolute count) vs. CD4+ cell density (cells/mm2) for tumor tissue. Pearson’s coefficient was used to analyze the association between the number of predicted pathogen specific TCRs and CD4+ cell density in tumor tissue for n=5 patients. 4E: Predicted pathogen TCRs (absolute count) vs. clonality for tumor tissue. Pearson’s coefficient was used to analyze the association between the number of predicted pathogen TCRs and clonality in tumor tissue for n=5 patients.4F: Predicted pathogen TCRs (absolute count) vs. CD8+ cell density (cells/mm2) for tumor tissue. Pearson’s coefficient was used to analyze the association between the number of predicted pathogen specific TCRs and CD8+ cell density in tumor tissue for n=5 patients.4G: PD-L1 score vs. proportion of predicted pathogen-specific TCRs. Pearson’s coefficient was used to analyze the association between PD-L1 score and proportion of predicted pathogen specific TCRs in tumor tissue for n=5 patients.

## Discussion

Checkpoint blockade immunotherapy reactivates T cells exhibiting a given phenotype at the systemic level, regardless of antigen-specificity[20]. Considering PD-1 is indiscriminately expressed on T cells following antigen exposure, reactivation of these T cells may be suboptimal[21]. Thus, a better understanding of the T cell repertoire in the context of the lungs is needed. Here, we performed TCR sequencing on a series of matched samples from 21 patients with early-stage NSCLC. To assess T cell infiltration across the lungs, we obtained samples from tumors, tumor edges, as well as 1cm, 2cm, 5cm, and 10cm stepping away from the tumor in addition to from matched peripheral blood.

Our spatial analysis of T cell infiltration allowed us to assess spatial T cell distribution across the lungs. Although tumors and adjacent lungs have previously been compared by us and others, such a dissection of the lung spatial environment and its T cell infiltrate and repertoire has not yet been undertaken[14]. Analysis of the homology in the T cell repertoire between the tumor and adjacent lung regions proved consistent with our prior findings. Indeed, MOI between the tumor and adjacent lung was ∼0.28, consistent with a prior study by our group demonstrating a median of ∼0.3 between tumors and adjacent lungs[14]. The same could be said for MOI between the tumor and peripheral blood, which was ∼0.18 in our study and ∼0.15 in the same prior study[14]. However, an important dimension not captured in our prior study was the inclusion of the tumor edge, which showed the highest homology with the tumor, highlighting the overall decreasing gradient in T cell homology from the tumor edge (MOI=0.36), to adjacent lung (0.28), and peripheral blood (0.18). MOI between tumor and tumor edge was also below what was observed in our prior analysis of intratumor heterogeneity (MOI=0.85), as should be expected[4]. The lower homology between the tumor and all regions of the adjacent lungs is suggestive of the potential presence of immune and T cell exclusion mechanisms within the tumor, which prevent T cell infiltration and could therefore explain the higher homology outside the tumor[22, 23].

Our analysis revealed increased T cell diversity and decreased T cell clonality in the tumor compared to the tumor edge and adjacent lungs. This supports prior studies by our group in larger cohorts with a lower spatial resolution[14]. Interestingly, by using GLIPH2.0[17], we demonstrate that regions with the highest clonality also present the lowest number of predicted pathogen-specific TCRs. This lack of predicted pathogen-specific TCRs could suggest a higher probability of infiltration with tumor-specific TCRs. Although this could be influenced by the increased diversity in pathogen TCR-rich regions, positive correlation between PD-L1 and T cell clonality may be suggestive of adaptive resistance induced by T cell activation and IFN-γ secretion, further supporting our hypothesis.

Our study does present certain limitations. First, despite our ability to reproduce several findings from prior studies, our study suffers from a limited sample size, which may have limited our ability to attain statistical significance in certain settings. However, our analysis of 143 samples provides an unprecedented high-resolution analysis of the T cell repertoire in the lungs of NSCLC patients. Second, our analysis of immune phenotypes was unfortunately limited by a lack of tissue availability and a restriction to formalin-fixed paraffin-embedded tissues, preventing us from performing any deeper phenotyping and/or tying TCR sequence to phenotype as has recently been done by others using single cell approaches[24]. Lastly, our analysis of antigen-specificity via GLIPH2.0 remains predictive based on *in silico* analyses and will need to be validated with fresh samples via functional assays, although these were unfortunately unavailable in this context. Nonetheless, our study provides critical information as to the spatial CD4/CD8 T cell composition in the lungs which has not been described by others to date. Overall, our study reveals the exclusion in T cells at play across the lungs of patients with NSCLC, as well as the unique T cell, PD-L1, and pathogen-specific T cell distribution patterns within these patients.

## Data Availability

The authors declare that the data supporting the findings of this study are available within the paper and its Supplementary materials. All data generated during this study are included in this published article and its supplementary information files. All data in this study are available from the corresponding author with a reasonable request.

## Declarations

### Ethics approval and consent to participate

The study was approved by the Ethics Committee of Second Xiangya Hospital Central South University (IRB: 2020084).

### Consent for publication

All authors have read and approved the article.

### Authors’ contributions

Qikang Hu, Jianjun Zhang, Xuefeng Xia, Xin Yi, Alexandre Reuben and Fenglei Yu: conception and design, acquisition of data, data analysis and interpretation, manuscript drafting, critical revision; statistical analyses. Meredith Frank, Liyan Ji, Qiongzhi He, Yingqian Zhang and Jianjun Zhang: manuscript drafting. Jianjun Zhang,Yin Yi, Alexandre Reuben and Fenglei Yu: conceived of the idea, supervised this project and revised the manuscript. Meredith Frank, Liyan Ji, Qiongzhi He, Yingqian Zhang, Alexandre Reuben, Muyun Peng, Xiaofeng Chen and Xuefeng Xia: acquisition of data, data analysis and interpretation. Qikang Hu, Yang Gao, Meredith Frank, Muyun Peng, Xiaofeng Chen, Liyan Ji: contributed reagents/materials/analysis tools. Fenglei Yu, Yang Gao: obtained funding.

## Acknowledgements

We thank SAN VALLEY DIAGNOSTICS and Servicebio Co. for the assistance with IHC experiments.

## List of Abbreviations

ACT: adoptive cell therapy
CDR3: complementarities determining region 3
ICB: immune checkpoint blockade
ICP: immune cells present
IMGT: ImMunoGeneTics
MOI: Morisita overlap index
NSCLC: non-small cell lung cancer
TIL: tumor infiltrating lymphocytes
TMB: Tumor mutational burden

## References

1 Al-Shahrabani F, Vallbohmer D, Angenendt S, Knoefel WT Surgical strategies in the therapy of non-small cell lung cancer. World J Clin Oncol 2014;5:595–603.

2 Siegel RL, Miller KD, Fuchs HE, Jemal A Cancer Statistics, 2021. CA Cancer J Clin 2021;71:7–33.

3 Gadgeel SM, Ramalingam SS, Kalemkerian GP Treatment of lung cancer. Radiol Clin North Am 2012;50:961–74.

4 Reuben A, Gittelman R, Gao J, Zhang J, Yusko EC, Wu CJ et al. TCR Repertoire Intratumor Heterogeneity in Localized Lung Adenocarcinomas: An Association with Predicted Neoantigen Heterogeneity and Postsurgical Recurrence. Cancer Discov 2017;7:1088–97.

5 Creelan BC, Wang C, Teer JK, Toloza EM, Yao J, Kim S et al. Tumor-infiltrating lymphocyte treatment for anti-PD-1-resistant metastatic lung cancer: a phase 1 trial. Nat Med 2021;27:1410–8.

6 Jiang Z, Zhou Y, Huang J A Combination of Biomarkers Predict Response to Immune Checkpoint Blockade Therapy in Non-Small Cell Lung Cancer. Front Immunol 2021;12:813331.

7 Lauss M, Donia M, Harbst K, Andersen R, Mitra S, Rosengren F et al. Mutational and putative neoantigen load predict clinical benefit of adoptive T cell therapy in melanoma. Nat Commun 2017;8:1738.

8 Jardim DL, Goodman A, de Melo Gagliato D, Kurzrock R The Challenges of Tumor Mutational Burden as an Immunotherapy Biomarker. Cancer Cell 2021;39:154–73.

9 Rizvi H, Sanchez-Vega F, La K, Chatila W, Jonsson P, Halpenny D et al. Molecular Determinants of Response to Anti-Programmed Cell Death (PD)-1 and Anti-Programmed Death-Ligand 1 (PD-L1) Blockade in Patients With Non-Small-Cell Lung Cancer Profiled With Targeted Next-Generation Sequencing. J Clin Oncol 2018;36:633–41.

10 Buttner R, Longshore JW, Lopez-Rios F, Merkelbach-Bruse S, Normanno N, Rouleau E et al. (2019) Implementing TMB measurement in clinical practice: considerations on assay requirements. ESMO Open, 2019/02/23 edn. pp. e000442

11 High TMB Predicts Immunotherapy Benefit. Cancer Discov 2018;8:668.

12 Kim SH, Go SI, Song DH, Park SW, Kim HR, Jang I et al. Prognostic impact of CD8 and programmed death-ligand 1 expression in patients with resectable non-small cell lung cancer. Br J Cancer 2019;120:547–54.

13 Feldmeyer L, Hudgens CW, Ray-Lyons G, Nagarajan P, Aung PP, Curry JL et al. Density, Distribution, and Composition of Immune Infiltrates Correlate with Survival in Merkel Cell Carcinoma. Clin Cancer Res 2016;22:5553–63.

14 Reuben A, Zhang J, Chiou SH, Gittelman RM, Li J, Lee WC et al. Comprehensive T cell repertoire characterization of non-small cell lung cancer. Nat Commun 2020;11:603.

15 Tian P, Zeng H, Ji L, Ding Z, Ren L, Gao W et al. Lung adenocarcinoma with ERBB2 exon 20 insertions: Comutations and immunogenomic features related to chemoimmunotherapy. Lung Cancer 2021;160:50–8.

16 Han J, Yu R, Duan J, Li J, Zhao W, Feng G et al. Weighting tumor-specific TCR repertoires as a classifier to stratify the immunotherapy delivery in non-small cell lung cancers. Sci Adv 2021;7.

17 Huang H, Wang C, Rubelt F, Scriba TJ, Davis MM Analyzing the Mycobacterium tuberculosis immune response by T-cell receptor clustering with GLIPH2 and genomewide antigen screening. Nat Biotechnol 2020;38:1194–202.

18 Simoni Y, Becht E, Fehlings M, Loh CY, Koo SL, Teng KWW et al. Bystander CD8(+) T cells are abundant and phenotypically distinct in human tumour infiltrates. Nature 2018;557:575–9.

19 Scheper W, Kelderman S, Fanchi LF, Linnemann C, Bendle G, de Rooij MAJ et al. Low and variable tumor reactivity of the intratumoral TCR repertoire in human cancers. Nat Med 2019;25:89–94.

20 Sharma P, Allison JP The future of immune checkpoint therapy. Science 2015;348:56–61.

21 Granier C, De Guillebon E, Blanc C, Roussel H, Badoual C, Colin E et al. Mechanisms of action and rationale for the use of checkpoint inhibitors in cancer. ESMO Open 2017;2:e000213.

22 Peranzoni E, Lemoine J, Vimeux L, Feuillet V, Barrin S, Kantari-Mimoun C et al. Macrophages impede CD8 T cells from reaching tumor cells and limit the efficacy of anti-PD-1 treatment. Proc Natl Acad Sci U S A 2018;115:E4041–e50.

23 Spranger S Mechanisms of tumor escape in the context of the T-cell-inflamed and the non-T-cell-inflamed tumor microenvironment. Int Immunol 2016;28:383–91.

24 Liu B, Hu X, Feng K, Gao R, Xue Z, Zhang S et al. Temporal single-cell tracing reveals clonal revival and expansion of precursor exhausted T cells during anti-PD-1 therapy in lung cancer. Nat Cancer 2022;3:108–21.

