## Supplementary Fig for "Spatial heterogeneity of T cell repertoire across NSCLC tumors, tumor edges, adjacent and distant lung tissues"

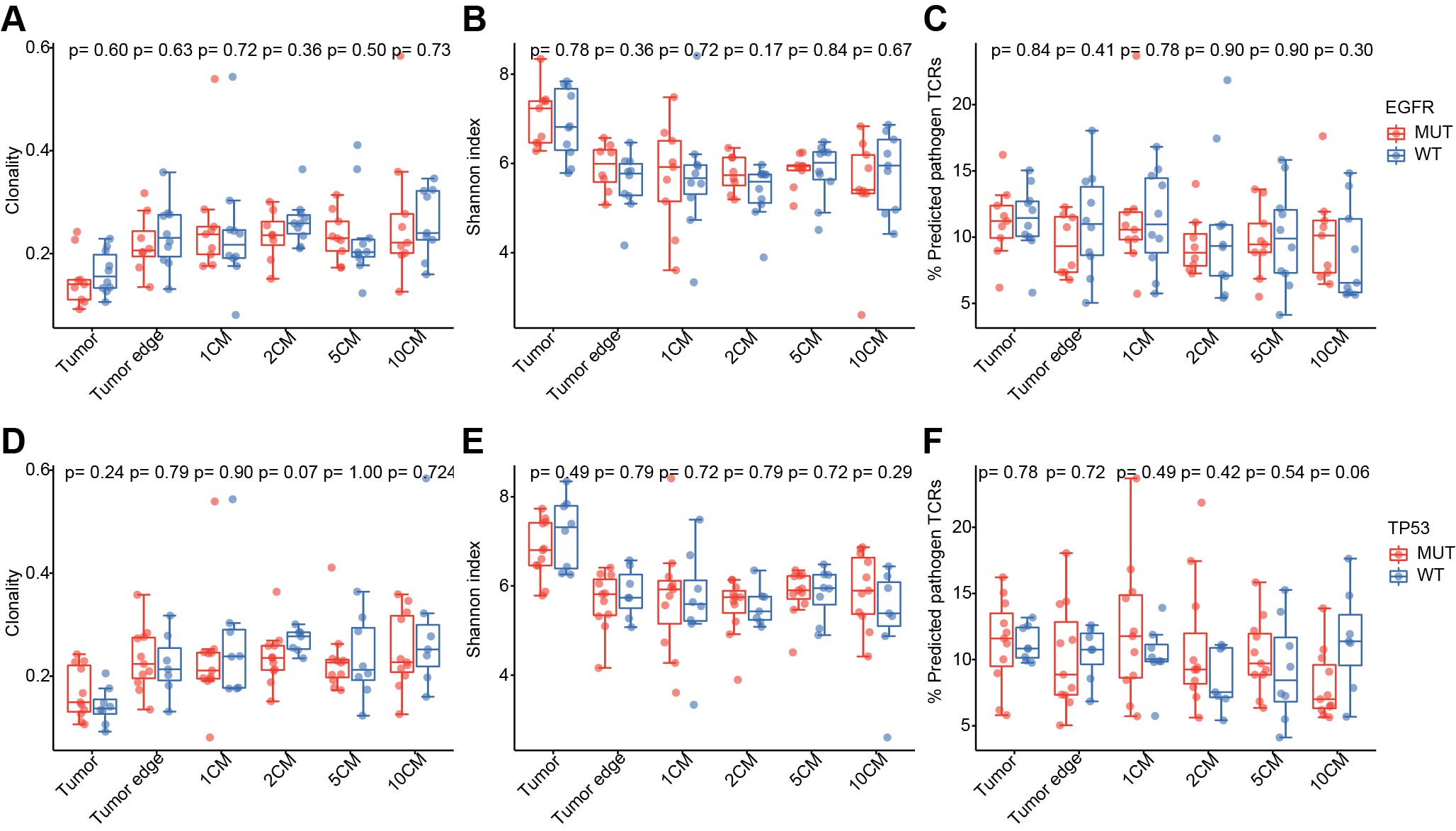

Supplementary Figure S1. Clonality (A), Shannon index (B), and percentage of predicted pathogen TCRs (C) were compared between wildtype tumors (WT) and tumors harboring mutations in EGFR across n=21 patients. Clonality (D), Shannon index (E), and percentage of predicted pathogen TCRs (F) were compared between wildtype tumors (WT) and tumors harboring mutations in GNAQ across n=21 patients.

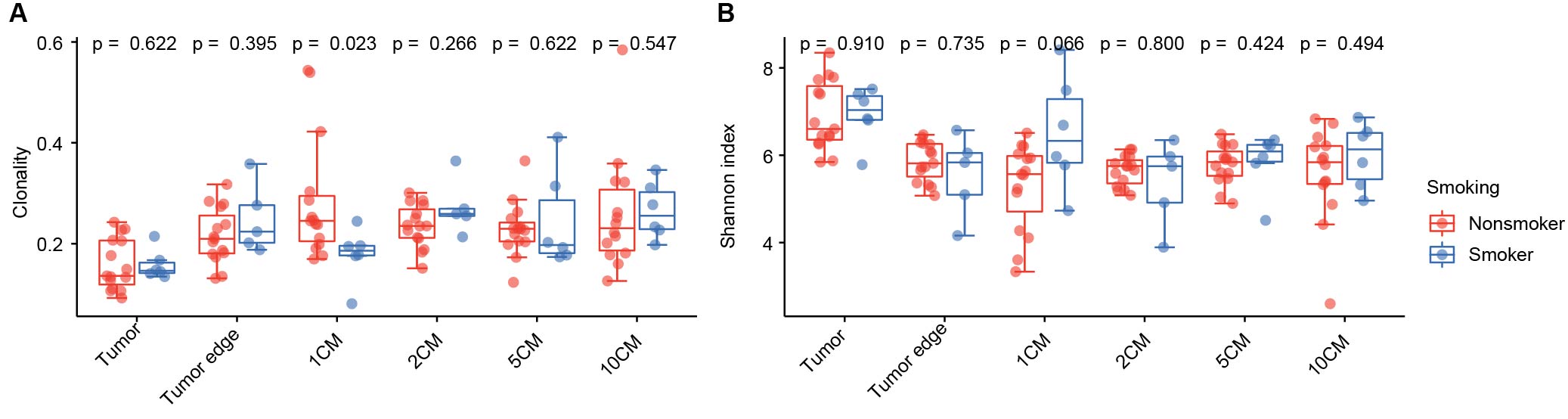

Supplementary Figure S2. (A) Median clonality values in smokers vs non-smokers across regions for n=21 patients. (B) Median Shannon index values in smokers vs non-smokers across regions for n=21 patients.

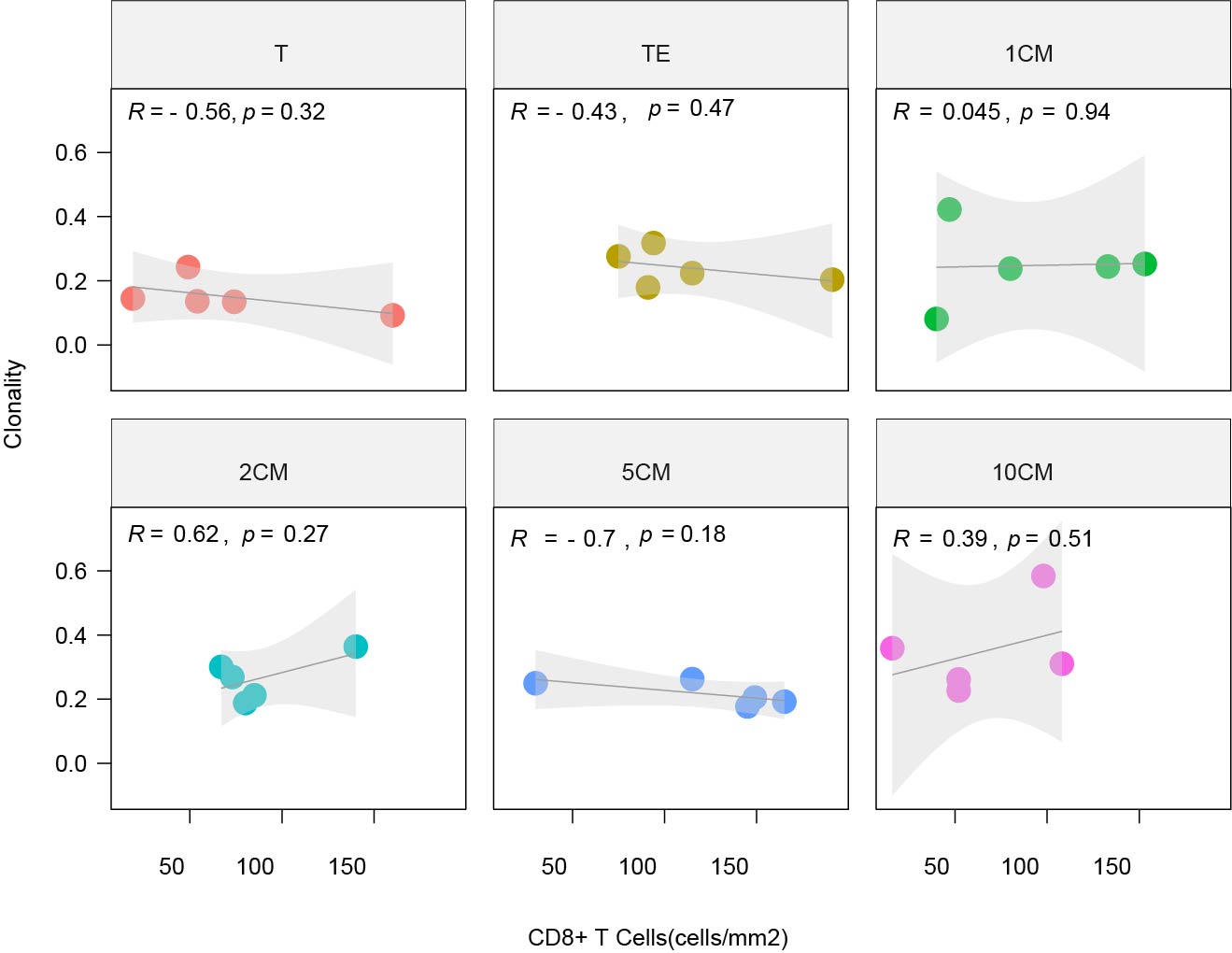

Supplementary Figure S3. Clonality vs CD8+ cell density (cells/mm2). Pearson’s coefficient was used to analyze the association between clonality and CD8+ cell density across all tissues for n=5 patients.

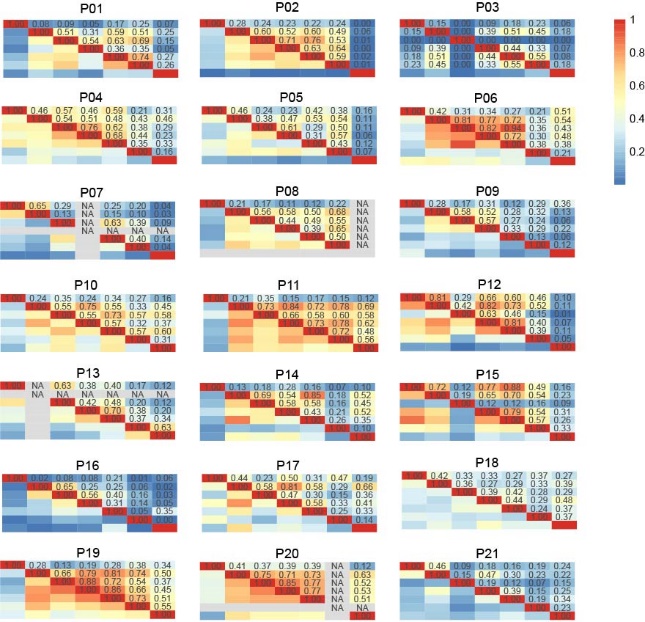

Supplementary Figure S4. MHoverlap for each patient.

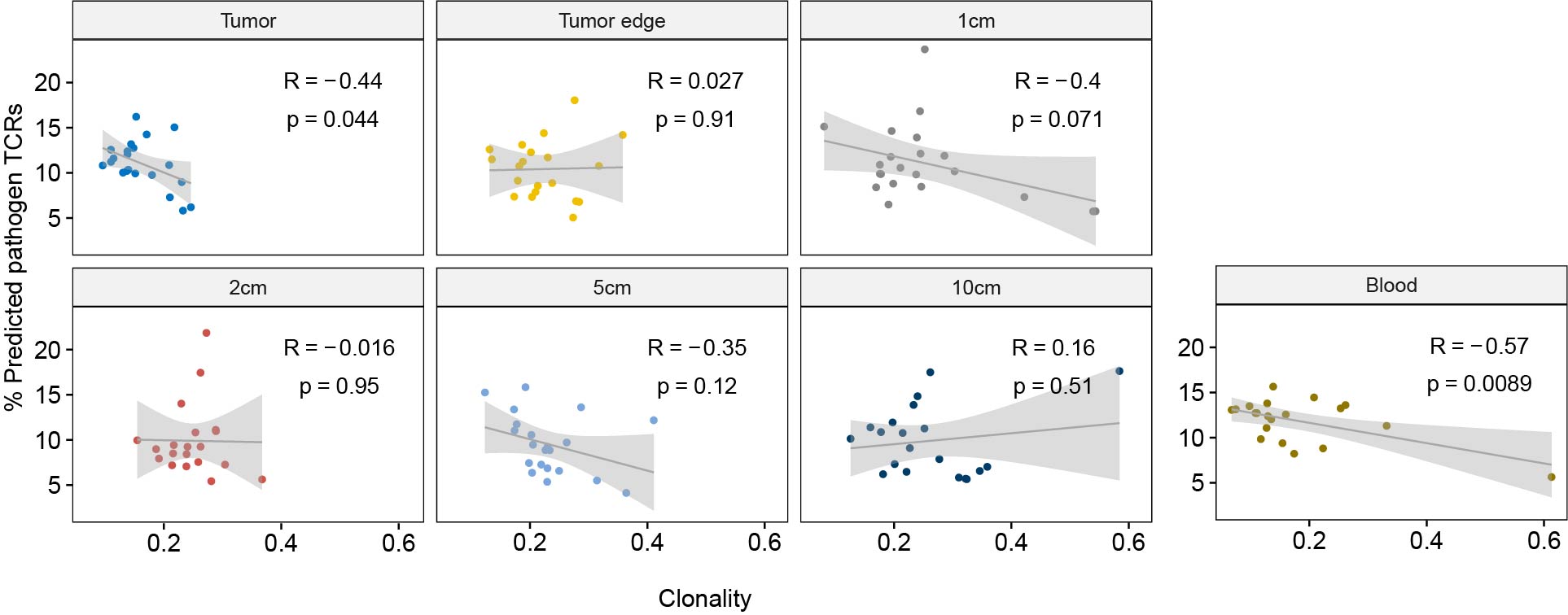

Supplementary Figure S5. Regional clonality values were plotted against median proportion of predicted pathogen TCRs for all tissues. The solid line represents correlation between clonality and predicted pathogen TCRs. Thin dotted lines represent the 95% confidence interval

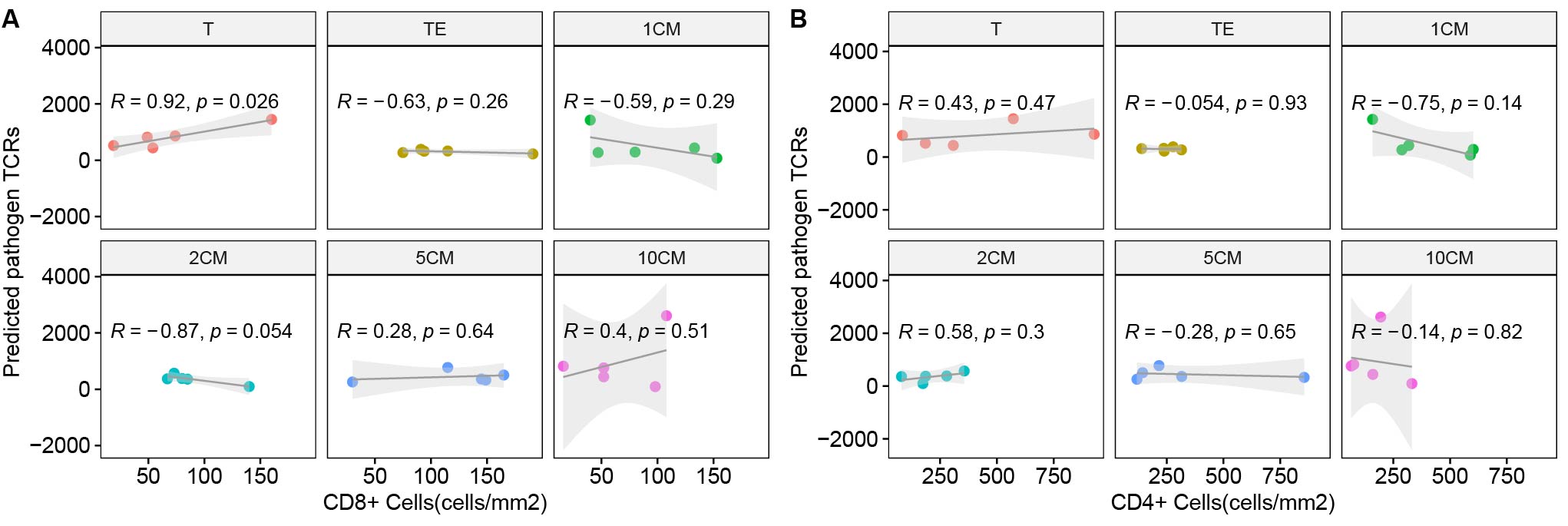

Supplementary Figure S6. (A) Number of predicted pathogen TCRs vs CD8+ cell density (cells/mm2). Pearson’s coefficient was used to analyze the association between number of predicted pathogen TCRs and CD8+ cell density across all tissues for n=5 patients. (B) Number of predicted pathogen TCRs vs CD4+ cell density (cells/mm2). Pearson’s coefficient was used to analyze the association between number of predicted pathogen TCRs and CD4+ cell density across all tissues for n=5 patients.
